# Modeling Clinical Trial Attrition Using Machine Intelligence: A driver analytics case study using 1,325 trials representing one million patients

**DOI:** 10.1101/2021.11.12.21266277

**Authors:** Emmette Hutchison, Youyi Zhang, Sreenath Nampally, Imran Khan Neelufer, Vlad Malkov, Jim Weatherall, Faisal Khan, Khader Shameer

**Author notes:** Equal contributors (Emmette Hutchison, Youyi Zhang, Sreenath Nampally).

## Abstract

The amount of time and resources invested in bringing novel therapeutics to market has increased year over year with fewer successful treatments reaching patients. In the lifecycle of drug development, the clinical phase is a major contributor to this decreasing efficiency in the development of clinical trials. One major barrier to the successful execution of a randomized control trial (RCT) is the attrition of patients who no longer participate in a trial either following enrollment or randomization. To address this problem, we have assembled a unique dataset by integrating multiple public databases including ClinicalTrials.gov and Aggregate Analysis of ClincalTrials.gov (AACT) to assemble a trial sponsor-independent dataset. This data spans 20 years of clinical trials and over 1 million patients (3,175 cohorts consisting of 1,020,085 patients and 79 curated features) in the respiratory domain and enabled a data-driven approach to identify top features influencing patient attrition in a trial. Top Features included *Duration of Trial, Duration of Treatment, Indication*, and *Number of Adverse Events*. We evaluated multiple machine learning models and found the best performance on the Test Set with Random Forest (Test subset: n=637 cohorts; RMSE 6.64). We envisage that our work will enable clinical trial sponsors to optimize trial run time by better anticipating and correcting for potential patient attrition using patient-centric strategies to improve patient engagement, thus enabling new therapies to be delivered to patients more quickly.

## Introduction

Drug development has become more costly with fewer successful new treatments brought to market year-after-year with one major contributor being the increasing time taken to complete clinical trials necessary to prove efficacy (Dickson & Gagnon, 2004; Scannell, Blanckley, Boldon, & Warrington, 2012). Large, randomized control trials (RCTs) in the clinical setting are complex endeavors that face many challenges to successful completion and determination of efficacy for a therapeutic agent. In addition to the efficacy of the agent being evaluated, as shown in Figure 1 successful completion of RCTs faces depend on many variables, including patient recruitment and patient retention that can delay the trial start, prolong the trial duration and result in failure to ascertain efficacy for the desired indication. (Stefan Harrer, Pratik Shah, Bhavna Antony, & Hu, 2019).

**Figure 1:**
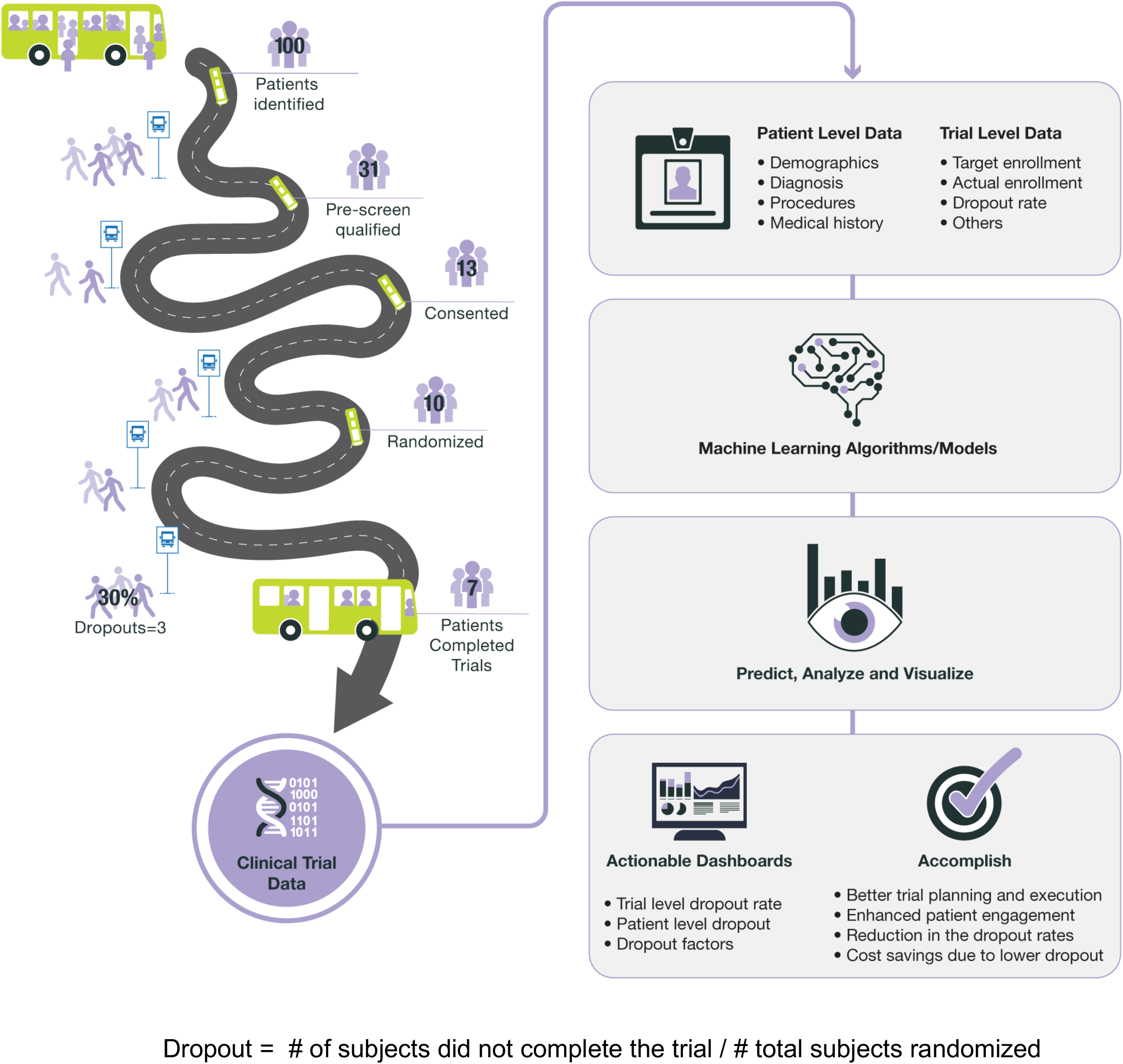
Overview of the problem of patient attrition in the clinical trial phase of drug development.

Patient attrition, also referred to as dropout or patient withdrawal, occurs when patients enrolled in a clinical trial either withdraw or are lost to follow-up by the clinical site and trial sponsor. Attrition can occur before trial randomization or after patients are randomized with the latter case having downstream effects that could lead to bias in trial populations where there is an interaction between a subpopulation and the treatment or protocol (Bell, Kenward, Fairclough, & Horton, 2013). Bias in trials can result in missing data and lead to failure to provide efficacy due to an insufficiently powered study, as has occurred in a few major cases. In one example from respiratory trial, COPD patients assigned to the placebo group withdrew due to an increased rate of adverse events (Burge et al., 2000; Jarad, Wedzicha, Burge, & Calverley, 1999).

Observed attrition rates in trials vary by indication, agent and trial protocol and can range up to 67% in the case of placebo groups in antipsychotic trials (Khan, Khan, Leventhal, & Brown, 2001). One review of dropout rates in trials published in leading journals found that 18% of trials surveyed had patient dropout rates above 20% (Wood, White, & Thompson, 2004). Drivers of attrition in previous analyses of clinical trials have been found to be specific to indication, population, quality of medical care and socioeconomic conditions. As one example, age of caretaker was one of the strongest observed predictors of patient dropout in pediatric asthma trials (Zebracki et al., 2003). Variability in patient attrition for a given trial context is poorly understood and it would be ideal for clinical trial sponsors to utilize a data-driven approach with past clinical trial data from different indications leveraged with techniques in the field of machine learning.

Recent years have seen rapid advances in machine learning and a general eagerness to adopt these methods in new domains, particularly in the clinical trials and biomedical space (Shah et al., 2019). Precision medicine in cardiology is being enabled through new applications of existing algorithms to imaging data and patient stratification (Johnson et al., 2017; Johnson et al., 2018). Recent work has even demonstrated automated interpretation of echocardiograms, raising the possibility of reducing clinician burden when interpreting this type of data (Ghorbani et al., 2020). Other efforts have developed imaging algorithms that can classify skin cancer with dermatologist-level performance and predict non-small-cell lung cancer (NSCLC) mutations from histopathological slides (Coudray et al., 2018). Machine learning models have also been developed to predict patient discharge diagnosis from electronic health record (EHR) data and leverage natural language processing (NLP) to interpret EHR-based radiology reports to predict oncologic outcomes (Kehl et al., 2019; Rajkomar et al., 2018). Could machine learning methodology be applied to the problem of patient dropout to yield clinical insight or improve trial optimization?

We propose a data-driven approach (see Figure 1) to addressing the problem of patient attrition using machine learning algorithms trained on publicly available industry sponsor-agnostic clinical trial data from the Aggregate Analysis of ClinicalTrials.gov (AACT) database sponsored by the Clinical Trials Transformation Initiative (CTT) (Alexander, Corrigan-Curay, & McClellan, 2018; Harrer, Shah, Antony, & Hu, 2019; Zarin, Tse, Williams, Califf, & Ide, 2011). The AACT database gives us the opportunity to investigate drivers of patient attrition in a sponsor agnostic manner and for the work presented in this study we have focused on respiratory studies.

## Results

### Assembly and characterization of a Respiratory Clinical Trials Dataset using public data from AACT

We first investigated the general characteristics of patient withdrawal in the AACT dataset in a sponsor agnostic manner across therapeutic areas (Figure 2A). The AACT database collects both aggregate data on individual clinical trials and also observations of deidentified, aggregated patient-level data associated with a given trial in the cohort or sub-group level. This observation-level data includes demographic information and adverse events. The largest fraction of patient drop-withdrawals in AACT was found in oncology trials with a 61% dropout rate observed followed by Renal trials (41%), Metabolic (31%), Immune (31%), Cardiovascular (23%) and Respiratory trials (23%). For our initial analysis, we chose to focus on building machine learning models of Respiratory trial patient attrition as, based on the prior literature of clinical trials in this therapeutic area, this area has been less well characterized. As such, we selected a cohort of cross-sponsor and cross-indication respiratory trials, see Figure 3A for the distribution of indications in this cohort. Overall, this dataset consisted of observations from 1,020,085 patients from 3,175 cohorts in 1,325 respiratory clinical trials.

**Figure 2:**
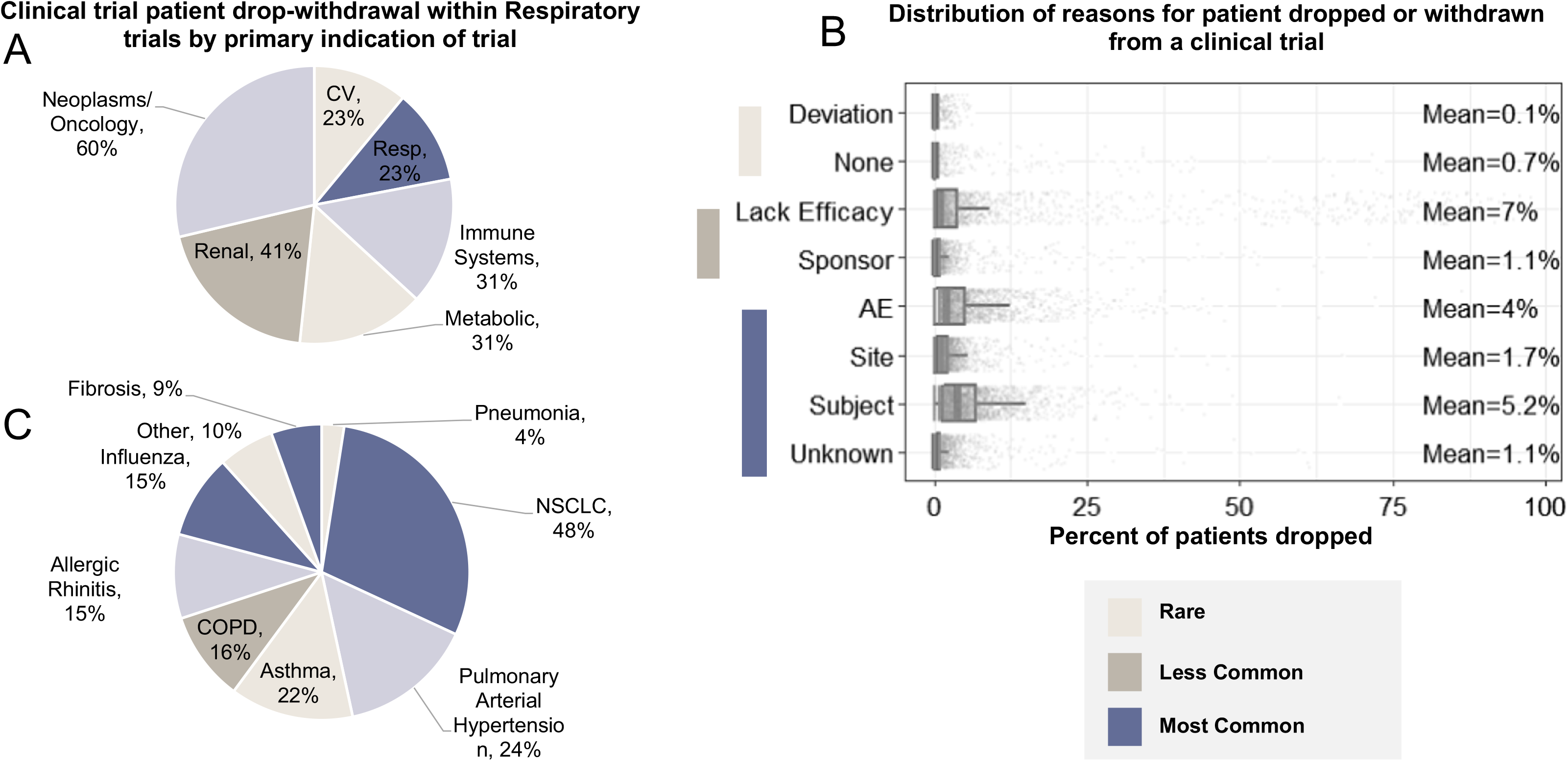
Industry-wide view of clinical trial patient drop/withdrawal reasons for the Respiratory Therapeutic Area. A. Clinical trial patient drop-withdrawal for all trials by therapeutic area of trial. B. Distribution of reasons for patient dropped or withdrawn from a clinical trial. C. Clinical trial patient drop-withdrawal within Respiratory trials by primary indication of trial. (N= 1,325)

**Figure 3:**
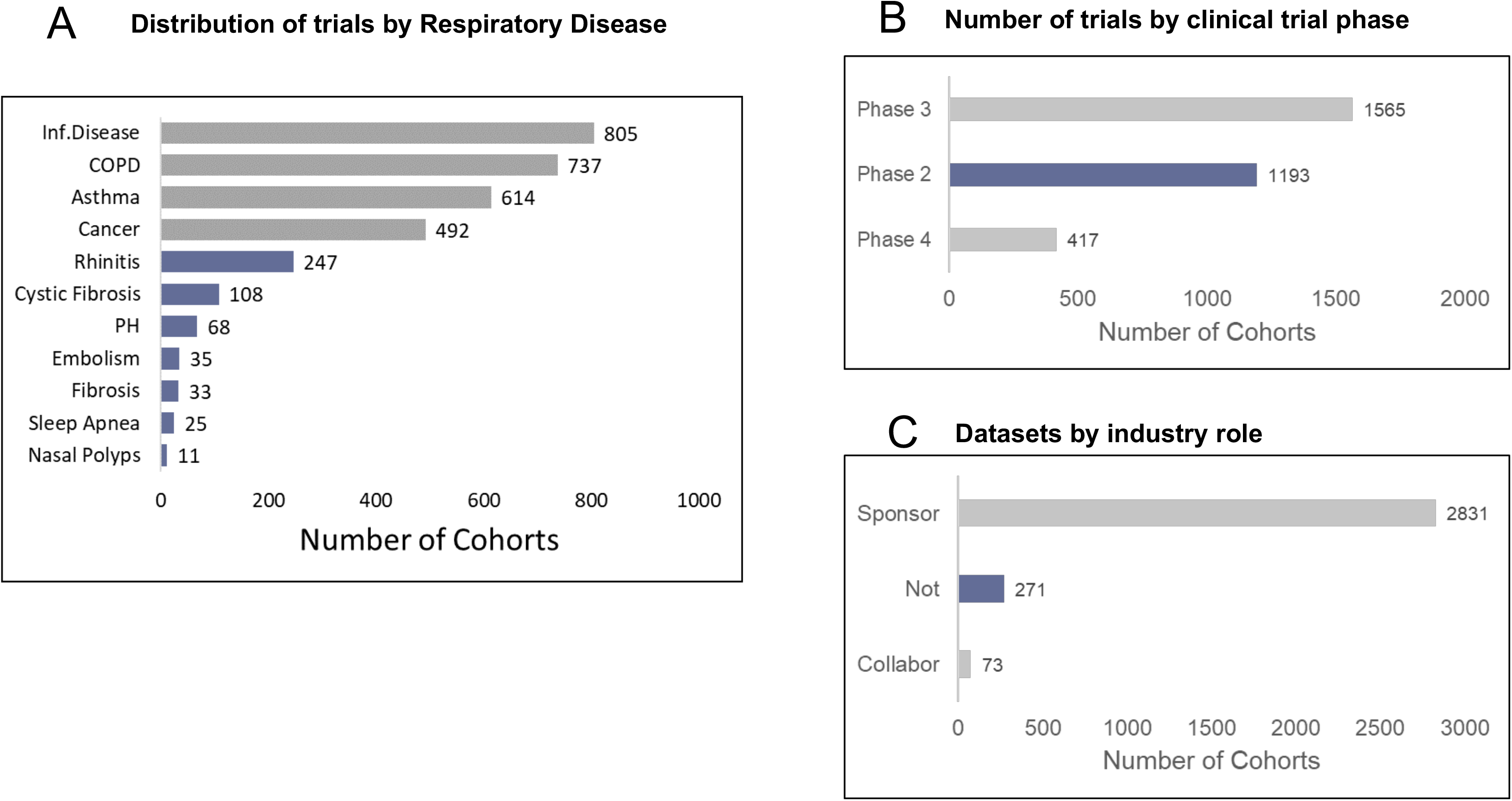
Characteristics of respiratory-focused AACT dataset for modeling patient attrition. A. Distribution of trials by Respiratory Disease. B. Number of trials by clinical trial phase. C. Datasets by industry role.

As we focused on respiratory trials, we next examined the distribution of reasons that a patient dropped out or was lost to follow-up for a trial (Figure 2B). Most frequently attributed reasons to a patient dropping out included “Unknown”, “Subject”, “Site” and “Adverse Event”. Less frequently observed reasons included “Deviation” and “None.” Lastly, we examined the distribution of indications within the Respiratory therapeutic area as this is the subset of AACT data that we considered for building a dataset from AACT and training machine learning models (Figure 2C). The remaining indications in the dataset with percentage of patients dropped were Pulmonary Arterial Hypertension (24%), Asthma (22%), COPD (16%), Allergic Rhinitis (15%), Influenza (15%), Other (10%), Fibrosis (9%) and Pneumonia (4%).

We next examined the distribution of trials in our respiratory trials dataset by indication (Figure 2). Trials selected from the AACT dataset over the past 20 years in the respiratory TA included 1,325 Clinical trials consisting of 3,175 cohorts. The majority of trials used for further analysis were for indications including Infectious Diseases (805 trials), COPD (737) and Asthma (614) as detailed in Figure 3A. The dataset assembled consisted of cohorts predominantly in Phase II (1193 cohorts) and Phase III (1565 cohorts) trials with 525 patient cohorts in Phase 4 respiratory trials included (Figure 3B). Of these patient cohorts, 89% were in trials run by a sponsor (Figure 3C). For a detailed overview of the assembly of the respiratory trials dataset, please see the end-to-end project workflow (Figure 4).

**Figure 4:**
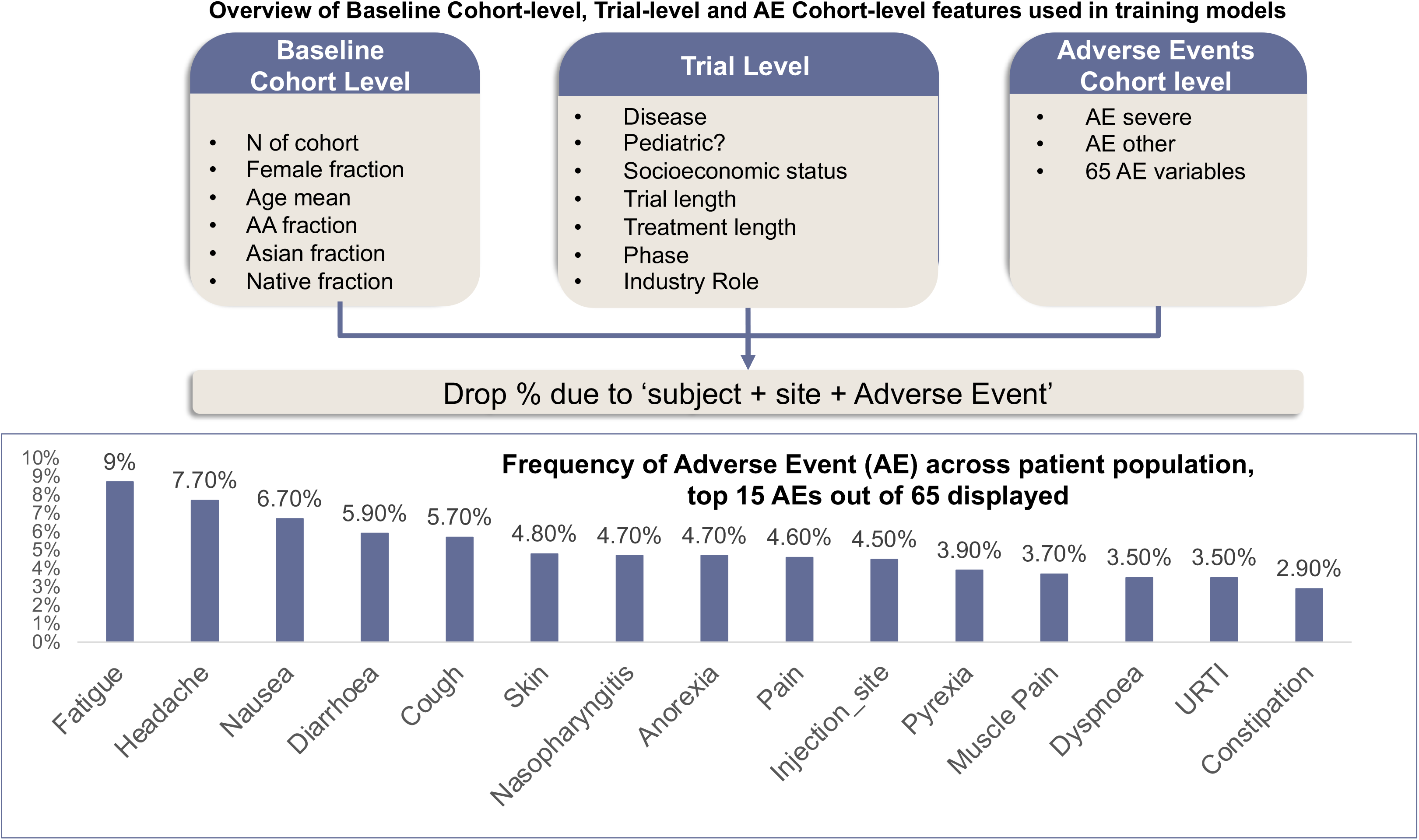
Workflow diagram for development of machine learning models of patient attrition based on AACT respiratory dataset.

### Characterization of Features from AACT Respiratory Clinical Trials Dataset using for Machine Learning

A total of 79 features were extracted from the dataset (Supplemental Table 1). Baseline Cohort-level Features included the cohort size, Female Fraction, Age Mean, African American Fraction, Asian Fraction and Native Fraction (includes American Indian or Alaska native and native Hawaiian or other pacific islander). Trial-level features included Disease, Pediatric Status, Socioeconomic Status, Trial Length, Treatment Length, Trial Phase and Industry Role. Adverse Events at the Cohort-Level include AE Severe, AE Other and 65 Adverse Event Features (see Top 15 AEs by frequency in Figure 5). The top 2 features, Duration of Trial and Duration of Treatment, were examined across indications and found to influence patient attrition variably across indication (Supplemental Figure 1). We noted that different disease indications have different patient attrition rate across clinical trials respiratory therapeutic area (Supplemental Figure 2 and 3). Fibrosis (18.99%), Sleep apnea (18.19) and cancer (16.21) trials were the indications with average dropout rate.

**Figure 5:**
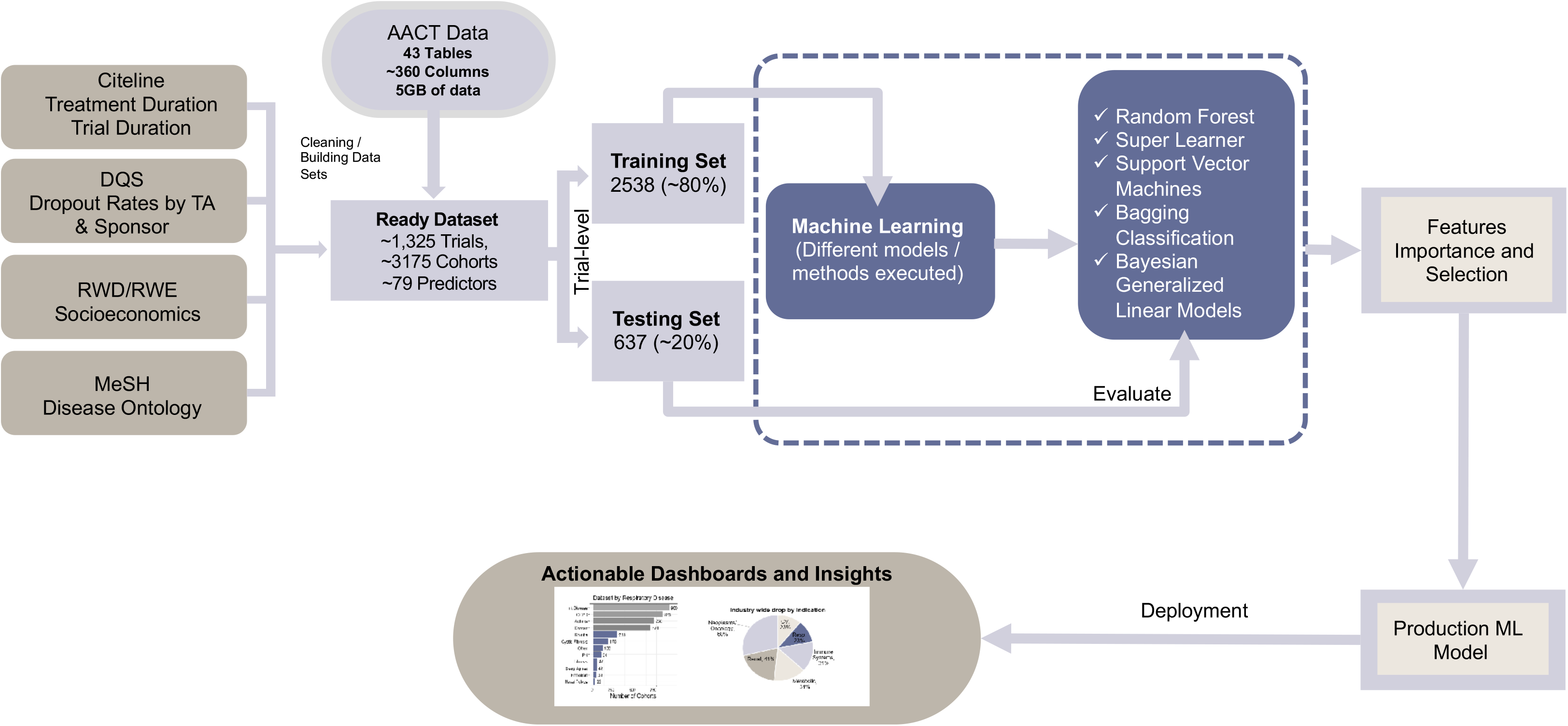
Features from AACT Respiratory Dataset Used for Development of Machine Learning Models. A. Overview of Baseline Cohort-level, Trial-level and AE Cohort-level features used in training models. B. Frequency of Adverse Event (AE) across patient population, top 15 AEs out of 65 displayed.

Recursive Feature Elimination (RFE) was performed to determine feature importance (Figure 6A). To determine the optimal number of features, RFE was performed with 10-fold cross validation and the optimal number of features evaluated by Root Mean Square Error (RMSE; Figure 6B) and Mean Absolute Error (MAE; Figure 6C). Based on this analysis, the top 8 features by RFE ranking were included in further analysis. These top features include Duration of Trial, Duration of Treatment, Disease, Serious AE Total, N Total, GDP Weighted, African American Fraction and Age Mean.

**Figure 6:**
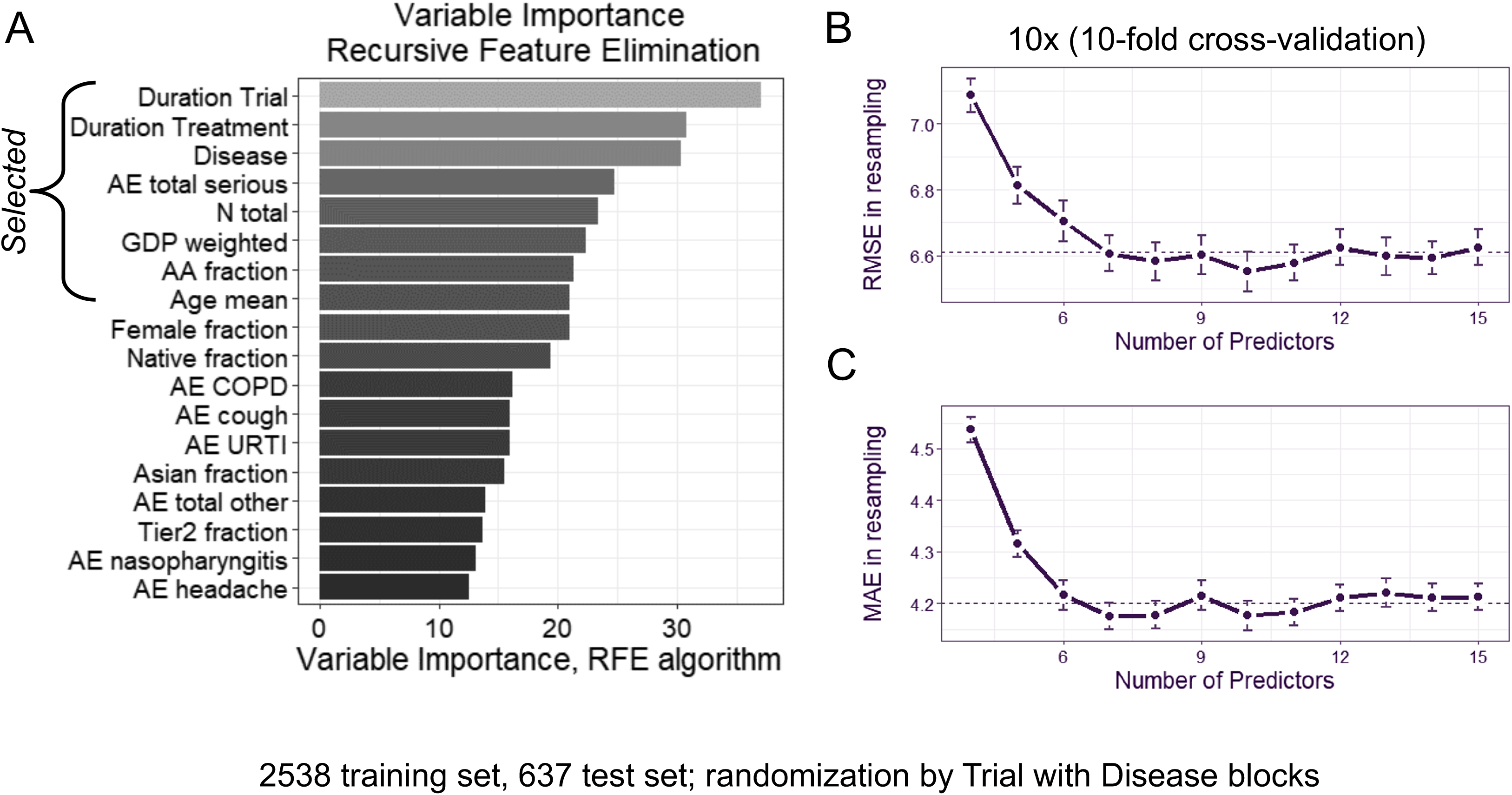
Models examined. (A) Variables ranked using Recursive Feature Elimination (RFE) with top 8 selected variables displayed. RMSE (B) and MAE (C) in analysis resampled by ranked features showing that there is no additional gain after the top 8 features.

**Figure 7:** Evaluation & selection of features. Table1. (A) Table of metrics for all models examined. RMSE (B). Table2. (A) Table of results for Random Forest and Linear models with 8 predictors.

### Training of Machine Learning Models and Selection of Best Model for Predicting Clinical Trial Attrition

Following selection of features, multiple machine learning models were trained to predict patient attrition (Table 1). Random Forest was the best performing model with a RMSE of 6.64 (see Table 1 for metrics for all models examined.) Other models tested included by order of performance Super Learner (an ensemble method unique to the SuperLearner R package, see Methods), Support Vector Machines, Bagging Regression, Bayesian Generalized Linear Models, Generalized Linear Models, Ordinary Least Squares and Generalized Linear Models via penalized maximum likelihood.

For thoroughness, we examined model performance of the top model on the Test set to determine that the top 8 Features determined by RFE generated optimal performance on the test set as well. In this context, Random Forest performance on the test set was evaluated by RMSE (Table 1). Model performance of Random Forest, the top model from SuperLearner, using the top 8 Features demonstrated that Random Forest was the optimal algorithm for predicting patient attrition with a RMSE of 6.25 (Table 2).

**Table 1:** Model performance. The super learner is an ensemble model, all estimates were run on 79 features.

| Algorithm | RMSE |
| --- | --- |
| Super Learner | 6.66 |
| Discrete Super Learner | 6.63 |
| Random Forest | 6.64 |
| Support Vector Machines | 7.29 |
| Bagging Classification | 7.70 |
| Bayesian Generalized Linear Models | 7.99 |
| Generalized Linear Models | 8.01 |
| Ordinary least squares | 8.01 |

**Table 2:** Table of results for Random Forest and Linear models with 8 predictors.

| Predictive Model | RMSE |
| --- | --- |
| Random Forest, 8 predictors | 6.25 |
| Linear Regression, 8 predictors | 8.37 |

## Discussion

This work assembled the first large-scale sponsor independent clinical trial dataset to develop a data-driven approach to elucidate the drivers of patient attrition in clinical trials. The advantage of clinical trial sponsors adopting this approach to modeling is that the development and deployment of such algorithms enable clinical trial sponsors to evaluate dropout rates for a given therapeutic area across the entire industry for a period of time without bias. Interestingly, features identified in this work correspond well to drivers of enrollment identified in prior work.

Patient attrition can occur pre-randomization (early attrition) and post-randomization with the setting of attrition having different impacts on the trial. Early attrition will result in a delay of randomization and trial start until sufficient numbers of patients are obtained to begin the trial. Late attrition can disrupt study outcomes through loss of statistical power, missing data and introduces the possibility of bias, either between treatment cohorts or through loss of heterogeneity in the participants for one group or another (Bell et al., 2013; Leon, Demirtas, & Hedeker, 2007). An excellent example of such bias from differential withdrawal has been observed in COPD placebo groups where the treatment group may experience symptomatic benefits (Vestbo et al., 2011). In the respiratory therapeutic area, this has been of particular issue for large COPD trials as the placebo cohorts were typically treated with corticosteroids prior to the trial (Calverley & Rennard, 2007).

One analysis of early versus late dropout reasons in two oncology studies found differential factors contributed to dropout based on when the patient withdrew (Siddiqi, Sikorskii, Given, & Given, 2008). Factors found to contribute to the early phase included minority race, screening duration and education where symptom severity and quality of life became factors driving late dropout. Another study of 18 oncology trials found that features predicting attrition included study duration, minority race, education and adverse events (Hui, Glitza, Chisholm, Yennu, & Bruera, 2013). Unfortunately, the available data for our analysis did not have educational or quality of life features and did not distinguish between early and late attrition. We did, however, find that both minority race and adverse events were top-ranked features used in the final and highest-performing model. It is likely that due to the loss of resolution on early relative to late trial attrition resulted in a feature set and model predicting both.

More interestingly is the fact that this work was able to demonstrate a performant model for patient attrition across trials and sponsors without a resolution on the trial sites. Prior work has identified clinical trial site performance as a major predictor of patient attrition, with wealthier countries demonstrating higher rates of dropout (Gheorghiade et al., 2014; Greene et al., 2018). Although this relationship has not been as explicit in respiratory studies as in recent heart failure trials, it is possible that the identification of country income-level as demonstrated by weighted Gross Domestic Product per capita (GDP PPP) is capturing this effect in a novel therapeutic area based on 20 years of sponsor agnostic clinical trial data.

The current approach to addressing attrition adopted by most sponsors is to inflate the initial cohort size using data from previous trials (Little et al., 2012). A more comprehensive data-driven approach using machine learning models trained on data from hundreds of clinical trials in a given therapeutic area provides a more principled approach to addressing attrition and better assessment of the necessary cohort composition to avoid bias.

Although the dataset and modeling in this work was limited to trials in the respiratory therapeutic area, there are other promising applications of this approach. Recent Heart Failure (HF) trials have demonstrated geographic heterogeneity and it will be interesting to see how well this approach generalizes across other cohorts in therapeutic areas covered by the AACT database. While there are highly contextual features that are not captured by our present dataset that have been demonstrated to predict dropout, such as treatment failure in COPD or age of caregiver in pediatric asthma trials, our model demonstrates good performance across respiratory cohorts (Musuamba et al., 2015; Robinson, Adair, Coffey, Harris, & Burnside, 2016). In many cases such niche features might be a barrier for a trial sponsor to accurately obtain due to variability across trials, making a performant model to predict patient attrition from more general trial features even more critical.

Patients may discontinue participating in a trial due to a variety of reasons. Patient attrition can be broadly classified as clinical (adverse events, injury, unrelated illness, comorbidities, etc.) personal (life-changing events like wedding, pregnancy etc.), or operational (unable to commute to the clinical trial sites, PI moved to another organization etc.) reasons. Understanding the precise reason for patient attrition and leveraging innovative strategies to use the information to improve patient engagement may lead to reduce the attrition rates. Further attrition factors could be incorporated into different clinical trial design scenarios including clinical trial forecasting, sample size estimation and planning. Such approaches would not only help to reduce the attrition but would help to address low Fragility Index (Tignanelli & Napolitano, 2019) of RCTs (Kipp, 2019).

The application of machine intelligence in the setting of drug discovery is attributed to an end-to-end transformation. However, large, enterprise-scale digital transformation is challenging and thus need a digital nudging approach. Where an individual process can be automated and transformed using intelligent automation and algorithms. While several works has been proposed to improve different facets of clinical trials using machine learning methods (Andrew, 2019; Lo, 2019; Lutz et al., 2018; Pedersen, Mansourvar, Sortso, & Schmidt, 2019; S & A, 2017; Vamathevan et al., 2019; Woo, 2019), our work is the only development of a machine learning model(Pedersen et al., 2019) for patient attrition developed using a large-scale sponsor agnostic dataset to date. Our data-driven approach represents a new approach to addressing one of the most critical barriers to drug development and bringing new therapeutics to market to address the patient’s needs. Application of this methodology to other therapeutic areas and deployment within the design of respiratory trials could lead to efficiencies that enable faster clinical trials and improved patient care.

## Methods

Data was collected from clinicaltrials.gov by filtering criteria - completed studies, respiratory diseases for phases 2, 3 and 4. We also filtered trials between 01/01/1998 to 12/31/2018 which resulted in 6039 trials. The AACT database provided aggregate information on patients who did not complete the trial and the reasons for their withdrawal. These trials identifiers were then matched with AACT database to derive the withdrawal reasons, which mainly related to subject, site and adverse related reasons. Similarly, age fraction, gender fraction and race were calculated from different tables within AACT database.

Disease category was extracted and then cleaned to get category we clubbed disease type for example: SCLC, Sarcoma etc was tagged as Cancer for further processing. Citeline data was also extracted in batches and compared with our data to realize that only 1679 trial identifiers could be found. The main imputations performed after merging Citeline data with AACT was imputing “median_female_fraction per disease age group”, “baseline counts”, “Female_fraction”, “Mean_Age_Female”. We imputed the median age per disease from the age group calculated by imputing 46 values missing age group data. Random forest was used to predict “Duration.Treatment” for imputation. “Duration.Enrollment” and “race” information had missing values; hence a Random forest model was used to predict these values. To address missing data in baseline final dataset columns of “Female fraction” - zero was introduced and “Age_mean” was introduced, race fractions variable with missing values was updated as zeros.

After an initial evaluation of the AACT dataset, our first application of machine learning algorithms to this dataset aimed at predicting patient attrition in clinical trials focused on the respiratory therapeutic area. Final data set was stored with 1,020,085 patients in 1,325 trials, 3,175 cohorts and 79 listed predictors. This dataset represents both cross-sponsor and cross-indication respiratory trials taking place over 20 years with both aggregate trial-level characteristics as well as patient-level observations regarding reasons for trial dropout or withdrawal. All analysis presented was at the cohort-level.

A total of over 3,175 completed respiratory cohorts were selected based on criteria including indication (see Figure 3A), Phase (Phase II to Phase IV; Figure 3B) and sponsor (Figure 3C).

Our analysis aimed to investigate multiple competitive machine learning algorithms in different settings, including an ensemble model that took optimal weighted averaged of our separate algorithms. Criteria to select machine learning models for further exploration depended on evaluation of model performance through cross-validation. Following cleaning and extraction, transformation and loading (ETL) of the dataset in the R programming language (see associated code and dataset artifacts), we split the entire data set into 80% and 20%, training and testing respectively. We then trained different machine learning models on our training set with parameter estimation based on 10-fold cross validation and gained our performance evaluation result on the testing set. Candidate models were considered into our modeling pool starting with our benchmark model Ordinary Least Squares as the baseline. To enhance our predictive accuracy as well as statistical interpretability, we also applied linear model with feature selection and regularization from either Bayesian or Frequentist Maximum likelihood perspective (Bayesian Generalized Linear Models, Support Vector Machines, Generalized Linear Models via penalized maximum likelihood). Besides typical regression-based models, we further explored tree-based approaches which have been shown to have better performance in the literature when dealing with large feature sets. In order to take the maximum use of the existing separate models, we applied aggregated algorithms such as Bagging, Super Learner model ensemble method. With all these algorithms implemented supported by SuperLearner (van der Laan, Polley, & Hubbard, 2007), we listed their performance based on RMSE values shown in Table 1, where we figured out that Random Forest had the potentiality towards further investigation.

To dig deeper into our random forest algorithm for performance enhancement, we implemented further hyperparameter tuning through grid search and feature selection via RFE. As random forest comprises multiple hyperparameters such as number of trees we used in the forest, maximum number of features considered for splitting, bootstrap strategy and etc., we implemented a grid search on the former two as the key ones that led to high impact of the model performance. Simultaneously, we applied RF-RFE (Granitto, Baiasioli, Furlanello, & Gasperi, 2006) which is a well-established feature selection method that is adapted to Random Forest, where it fits the algorithm and removes the weakest features iteratively until the performance metric reaches its optimal point.

## Data Availability

Dataset, data dictionary, and code of model development and feature selection is available at the GitHub URL: https://github.com/AstraZeneca/CTELC-Patient-Attrition-Model

## Competing Interests

All authors were employees of AstraZeneca at the time of the execution of this work.

## Acknowledgements

Authors would like to thank Dr. Christopher Miller, and Rosa Lamarca for their help with the data preparation.

## Author Contributions

EH compiled a first draft of the manuscript with contributions from all coauthors. YZ, SN, INK performed the analyses. VM compiled an initial version of the data and model. SN and INK compiled the final version of the data. KS designed the study and supervised the data science team. FK and JW provided critical feedback and reviewed the manuscript.

## Figures and Legends

Supplemental Figure 1. Respiratory trails from AACT. (A) Effect of Trial Duration and Duration of Treatment, Drop withdrawal by disease (B) by disease –, Longer Trials and Longer Treatment cause higher Patient Attrition. It also differs by Disease, i.e. higher in Cancer, Fibrosis, Pulmonary Hypertension

Supplemental Figure 2. Indications associated with average patient attrition rates across the respiratory therapeutic area

Supplemental Figure 3. Box plots of 11 indications and percentage level attrition rates compiled from the study cohort

Supplemental Table 1. List of 79 features and their importance based on mean decrease in accuracy.

