## Supplemental for "Modeling Clinical Trial Attrition Using Machine Intelligence: A driver analytics case study using 1,325 trials representing one million patients"

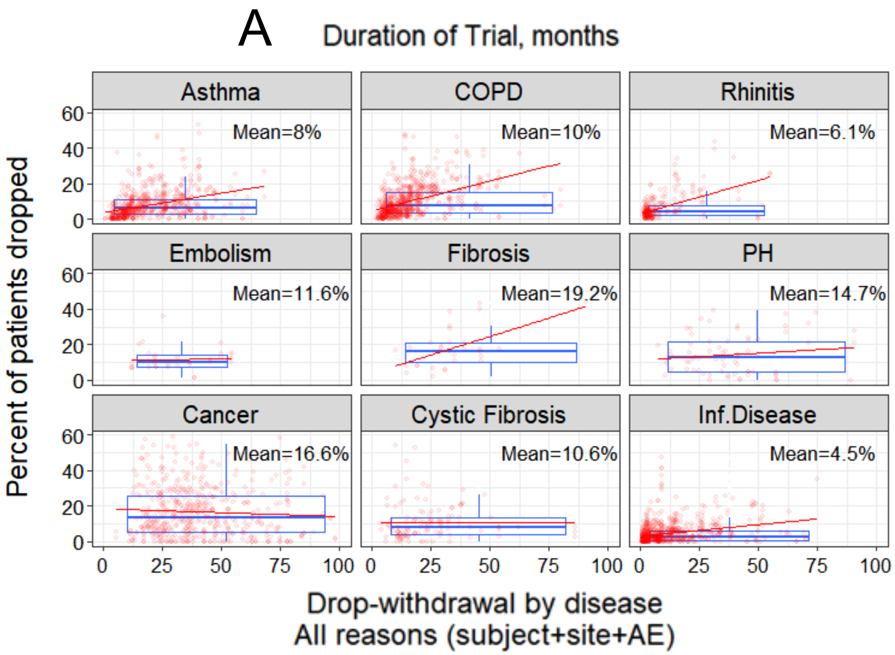


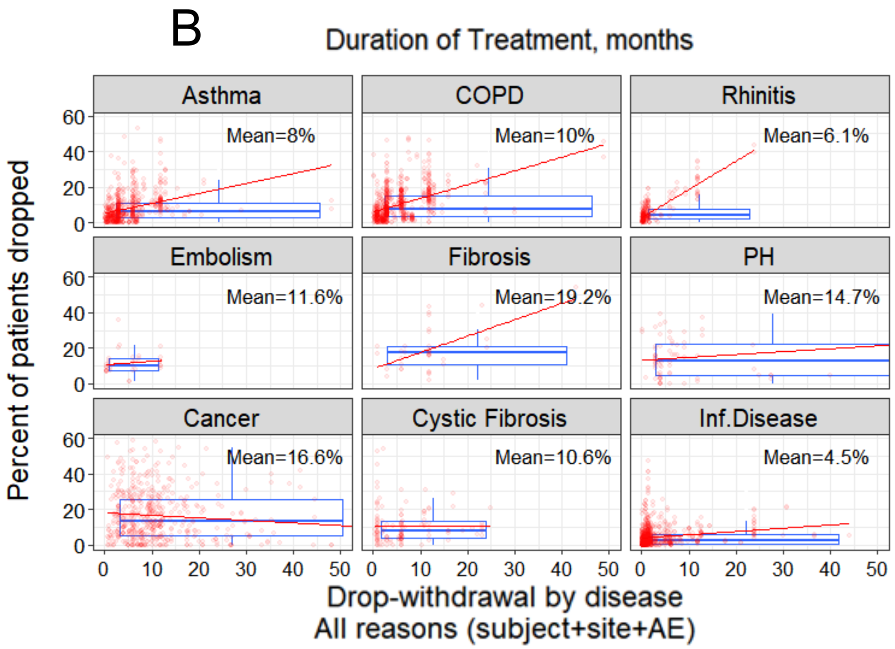


Supplemental Figure 1

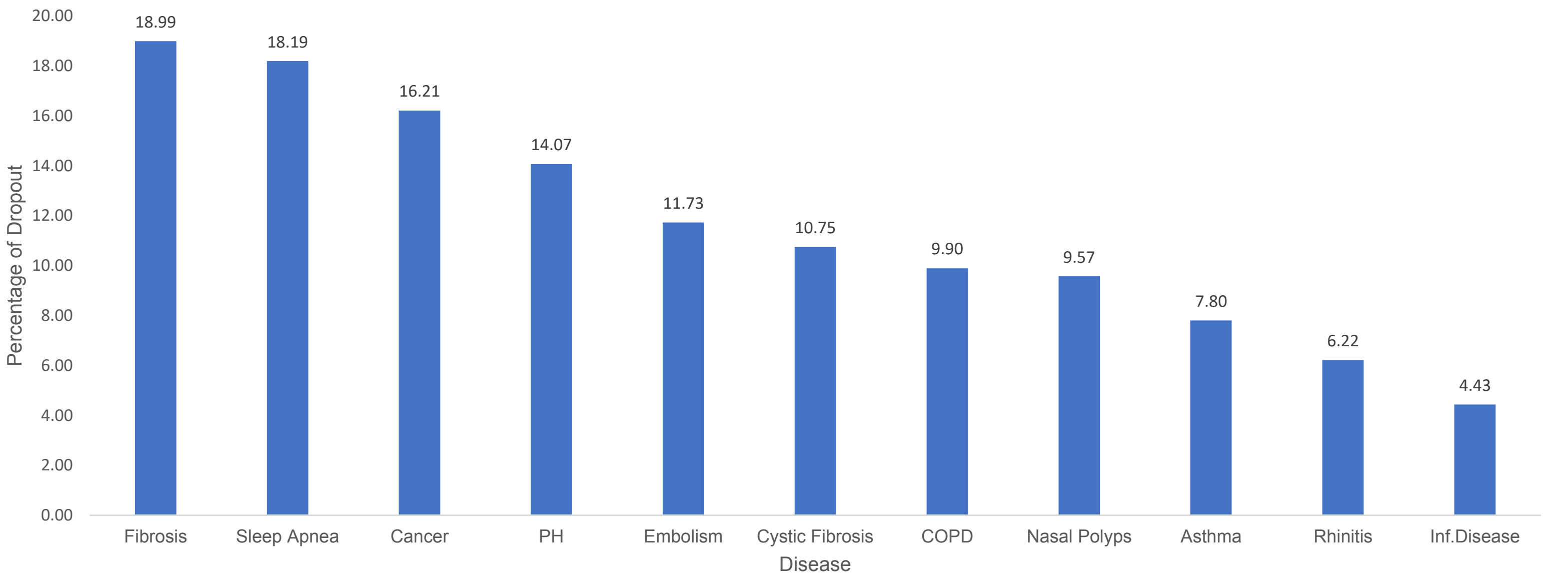


Supplemental Figure 2

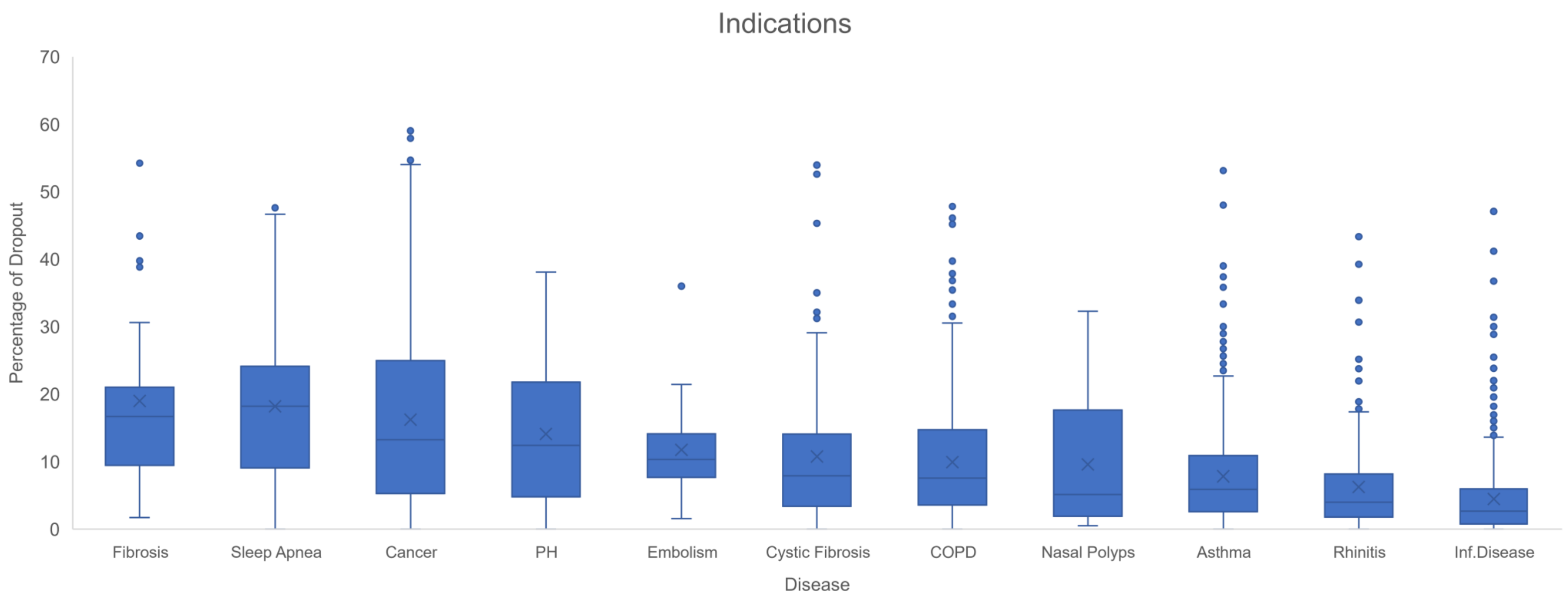


Supplemental Figure 3

| Variable Name | Importance | Variable Name | Importance | Variable Name | Importance | Variable Name | Importance |
|---|---|---|---|---|---|---|---|
| Duration.Trial | 3.71400 | AE_URTI | 0.70441 | AE_chest_pain | 0.38369 | AE_drowsiness | 0.20765 |
| Duration.Treatment | 2.42106 | AE_diarrhoea | 0.69946 | Age_mean | 0.38151 | Tier2_fraction | 0.20272 |
| AE_total_serious | 1.61331 | AE_hypertension | 0.66051 | AE_chills | 0.37708 | AE_swelling | 0.19447 |
| Disease_n | 1.57841 | AE_abdominal_pain | 0.64610 | AE_syncope | 0.37430 | AE_stroke | 0.16498 |
| AE_respiratory_failure | 1.54807 | AE_bleeding | 0.63590 | AE_insomnia | 0.36911 | AE_hypotension | 0.14925 |
| AE_nausea | 1.49477 | AE_neutropenia | 0.61036 | AE_heart_attack | 0.35395 | AE_thrombocytopenia | 0.14853 |
| AE_back_pain | 1.34323 | AE_AST_ALT | 0.59867 | AE_asthma | 0.34533 | AE_embolism | 0.14507 |
| AE_nasopharyngitis | 1.32966 | AE_constipation | 0.58980 | Phase_n | 0.30753 | AE_skin | 0.14350 |
| AE_dyspnoea | 1.27339 | AE_hyponatraemia | 0.55089 | AE_pneumothorax | 0.30710 | AE_nasal | 0.12929 |
| GDP_weighted | 1.08668 | Native_fraction | 0.50362 | AE_heart_failure | 0.30395 | AE_alopecia | 0.12598 |
| AE_COPD | 1.00711 | AE_pain | 0.50243 | AE_GORD | 0.30071 | AE_thrombosis | 0.10139 |
| AE_UTI | 0.99751 | AE_dehydration | 0.49400 | AE_LRTI | 0.29124 | AE_hyperglycaemia | 0.09024 |
| AE_cough | 0.98649 | AE_muscle_pain | 0.49087 | AE_mood | 0.28042 | Industry_n | 0.08609 |
| Female_fraction | 0.92635 | AE_injection_site | 0.41509 | AE_joint_pain | 0.27656 | AE_fibrillation | 0.08405 |
| AE_total_other | 0.92028 | Tier3_fraction | 0.41373 | AE_dyspepsia | 0.27238 | AE_pleural_effusion | 0.07435 |
| AE_fatigue | 0.84326 | AE_anorexia | 0.41210 | AE_renal_failure | 0.22347 | AE_hypoxia | 0.05447 |
| AA_fraction | 0.80997 | AE_limb_pain | 0.40129 | AE_gastroenteritis | 0.21740 | AE_total_mortality | 0.04156 |
| AE_hypokalaemia | 0.79294 | AE_headache | 0.39666 | AE_haemoptysis | 0.21384 | AE_taste | 0.02010 |
| AE_dizziness | 0.78284 | AE_sepsis | 0.39120 | Asian_fraction | 0.21010 | AE_pharynx_pain | 0.00119 |
| N_total | 0.74820 | AE_pyrexia | 0.38522 | AE_anaemia | 0.20896 | | |

Supplemental Table 1
